# SARS-CoV-2 B.1.1.7 lineage-related perceptions, COVID-19 vaccine acceptance and travel worry among healthcare workers

**DOI:** 10.1101/2021.01.19.21250111

**Authors:** Mohamad-Hani Temsah, Mazin Barry, Fadi Aljamaan, Abdullah N Alhuzaimi, Ayman Al-Eyadhy, Basema Saddik, Fahad Alsohime, Ali Alhaboob, Khalid Alhasan, Ali Alaraj, Rabih Halwani, Amr Jamal, Nurah Alamro, Reem Temsah, Samia A. Esmaeil, Shelaweeh Alanazi, Fahad Alzamil, Ali Alsomaily, Jafar A. Al-Tawfiq

## Abstract

**Background:** Healthcare workers’ (HCWs’) travel-related anxiety needs to be assessed in light of the emergence of SARS-CoV-2 mutations.

**Methods:** An online, cross-sectional questionnaire among HCWs between December 21, 2020 to January 7, 2021. The outcome variables were HCWs’ knowledge and awareness of the SARS-CoV-2 B.1.1.7 lineage, and its associated travel worry and Generalized Anxiety Disorder (GAD-7) score.

**Results:** A total of 1,058 HCWs completed the survey; 66.5% were female, 59.0% were nurses. 9.0% indicated they had been previously diagnosed with COVID-19. Regarding the B.1.1.7 lineage, almost all (97.3%) were aware of its emergence, 73.8% were aware that it is more infectious, 78.0% thought it causes more severe disease, and only 50.0% knew that current COVID-19 vaccines are effective in preventing it. Despite this, 66.7% of HCWs were not registered to receive the vaccine. HCWs’ most common source of information about the new variant was social media platforms (67%), and this subgroup was significantly more worried about traveling. Nurses were more worried than physicians (P=0.001).

**Conclusions:** Most HCWs were aware of the emergence of SARS-CoV-2 B.1.1.7 variant and expressed substantial travel worries. Increased worry levels were found among HCWs who used social media as their main source of information, those with lower levels of COVID-19 vaccine uptake, and those with higher GAD-7 scores. The utilization of official social media platforms could improve accurate information dissemination among HCWs regarding the pandemic’s evolving mutations. Targeted vaccine campaigns are warranted to assure HCWs about the efficacy of COVID-19 vaccines toward SARS-CoV-2 variants.

## 1 Introduction

The emergence of the Severe Acute Respiratory Syndrome Coronavirus 2 (SARS-CoV-2) has resulted in a global pandemic. Being an RNA virus, SARS-CoV-2 can mutate over time, and thus multiple SARS-CoV-2 variants are circulating globally ^1^. As of December 13, 2020, there were 1,108 reported cases of people infected with the B.1.1.7 variant in the United Kingdom (UK) in almost 60 different local authorities, with the exact number likely much higher ^2^. The variant under investigation in this study was discovered in December 2020 (VUI-202012/01) and is characterized by a set of 17 mutations ^2^. One of the most significant mutations is N501Y in the gene coding of the spike protein that is used to attach to angiotensin-converting enzyme 2 receptors ^2^.

The B.1.1.7 lineage was first identified by the COVID-19 Genomics UK consortium, which performs random genetic sequencing of positive COVID-19 samples in the UK. Since its creation in April 2020, the consortium has sequenced 140,000 virus genomes from individuals infected with COVID-19 and publishes weekly reports on its website ^3^. This B.1.1.7 variant is estimated to have first emerged in the UK in September 2020 ^1^. As of December 26, 2020, more than 3,000 cases of this new variant have been reported in the UK^4^.

This mutation has been found to have a high transmission rate and has become one of the main circulating variants in several locations in the UK ^2^. This variant has since been detected in many other countries around the world, including the United States (U.S.) and Canada ^1^. The European Centre for Disease Prevention and Control (ECDC) recommends that residents of the most affected areas restrict movement and travel, including international travel outside of these areas ^4^. Several countries, including the UK, have imposed travel restrictions to limit the variant’s rapid spread ^4^.

Scientists and researchers have begun to learn more about this variant to better understand its virulence and infectiousness and whether currently authorized COVID-19 vaccines would illicit immunity against it ^1^. The rapid spread of B.1.1.7 has prompted the ECDC to announce that the overall risk associated with the introduction and further spread of this SARS-CoV-2 strain is high ^4^. Despite the current lack of evidence that these variants may cause a more severe illness or increased risk of death, healthcare workers (HCWs) are yet again challenged by a new stressful situation that could affect their psychological well-being and/or travel arrangements ^1,5^.

In the Kingdom of Saudi Arabia (KSA), the rapid spread of this new strain has prompted stricter international travel bans to and from affected countries (from December 21, 2020 to January 7, 2021). Media outlets have announced that many non-Saudi expat HCWs have been stranded abroad and are still struggling to return to the KSA, with available flights in short supply ^6^. Starting on December 17, 2020, Saudi Arabia has rolled out mass BNT162b2 vaccinations for HCWs, with open registration beginning a week earlier ^7^. As the rapid evolution of the situation warrants further research, we conducted this study among HCWs in Saudi Arabia to assess their perceptions, stress levels, and travel worries caused by this new SARS-COV-2 variant.

## 2 Method

### 2.1 Data Collection

This national cross-sectional survey was conducted among HCWs in Saudi Arabia during the COVID-19 pandemic. Data were collected between December 27, 2020 and January 3, 2021. At the time of data collection, there were at least a handful of countries that had reported infection with the B.1.1.7 variant of SARS-CoV-2. HCWs were surveyed regarding their perceptions, stress levels, and travel worry caused by the new variant. Participants were invited using a convenience sampling technique. We used several social media platforms and email lists to recruit participants. The survey was a pilot-validated, self-administered questionnaire that was sent to HCWs online through SurveyMonkey^©^, a platform that allows researchers to deploy and analyze surveys via the web. The questionnaire was adapted from our previously published study with modification and additions related to the new SARS-COV-2 variant ^8-10^.

The questions asked about respondents’ demographic characteristics (job category, age, sex, years of clinical experience, and work area), previous exposure to COVID-19 patients, travel history in the previous 3 months, and whether they had received and/or registered to receive the COVID-19 vaccine. We assessed the following outcomes related to the new SARS-COV-2 variant: knowledge, perceptions, and travel worries. In addition, we assessed factors affecting respondents’ worry level regarding international travel as well as HCWs’ sources of information about the B.1.1.7 SARS-CoV-2 mutant variant. HCWs’ anxiety was also measured by the validated 7-item General Anxiety Disorder (GAD-7) questionnaire, which has been used in several studies assessing HCWs’ anxiety levels during the pandemic ^9,11^.

HCWs were informed of the purpose of the study in English at the beginning of the online survey. The respondents were given the opportunity to ask questions via a dedicated email address for the study. The Institutional Review Board at the College of Medicine and King Saud University Medical City approved the study (approval #20/0065/IRB). A waiver for signed consent was obtained since the survey presented no more than a minimal risk to subjects and involved no procedures for which written consent is usually required. To maximize confidentiality, personal identifiers were not required.

### 2.2 Statistical analyses

Descriptive analyses using means and standard deviations were applied to continuously measured variables, and categorically measured variables were described with frequencies and percentages. The histogram and the statistical Kolmogorov–Smirnov tests of normality were used to assess the statistical normality of the measured continuous variables. The multiple response dichotomy analysis was applied to the multiple option questions, such as the one asking about sources of information. Respondents’ awareness of the new mutagenic SARS-COV-2 virus strain was measured with eight questions, which received a score of 1 for each correctly answered knowledge/awareness question and 0 for incorrectly answered questions. A total awareness of mutagenic viral outbreak score was measured by adding the scores for the knowledge indicators (range: 0–8 points). For categorically measured variables, the independent samples t-test and one-way ANOVA test were used to assess the statistical significance of HCWs’ mean perceived worry regarding travel. Pearson’s bivariate test of correlation was used to assess the correlations between metric variables. Multivariate linear regression analysis was used to assess the multivariate associations of HCWs’ demographic and professional characteristics and their perceptions with their worry level about travelling abroad. IBM SPSS Statistics for Windows, Version 21.0. Armonk, NY: IBM Corp. was used for the statistical data analysis, and results were considered significant at the 0.05 level.

## 3 Results

In total, 1,212 HCWs participated in the survey. Of these, 1,058 completed the survey, resulting in a completion rate of 87.2%. Most participants were female (66.5%), between the ages of 31 and 40 years (44.5%) and married (75.7%). Most respondents were nurses (59.2%), followed by physicians (38%). Participants worked in intensive care units (ICUs) (25.8%), general wards (24.7%), or outpatient departments (OPDs) (21.3%) in various hospital settings (Table 1).

**Table 1:** Descriptive analysis of HCWs’ sociodemographic and professional characteristics (N = 1058)

| Characteristic | n | % |
| --- | --- | --- |
| **Sex** |  |  |
| Male | 354 | 33.5 |
| Female | 704 | 66.5 |
| **Age** |  |  |
| 20–30 years | 238 | 22.5 |
| 31–40 years | 471 | 44.5 |
| 41–50 years | 263 | 24.9 |
| ≥ 51 years | 86 | 8.1 |
| **Marital status** |  |  |
| Single | 257 | 24.3 |
| Married | 801 | 75.7 |
| **Nationality** |  |  |
| Expatriate | 736 | 69.6 |
| Saudi | 322 | 30.4 |
| **Clinical role** |  |  |
| Consultant | 213 | 20.1 |
| Assistant consultant/fellow | 52 | 4.9 |
| Resident/registrar | 138 | 13.0 |
| Nurse | 626 | 59.2 |
| Intern/medical student | 29 | 2.7 |
| **Hospital ward** |  |  |
| ICU | 273 | 25.8 |
| ER | 110 | 10.4 |
| OR | 62 | 5.9 |
| Isolation ward | 63 | 6.0 |
| General ward | 261 | 24.7 |
| OPD | 225 | 21.3 |
| Non-clinical area | 64 | 6.0 |
| **Hospital sector** |  |  |
| Private | 174 | 16.4 |
| Public/governmental | 487 | 46.0 |
| University hospital | 397 | 37.5 |
| **Hospital specialty** |  |  |
| Primary healthcare center | 123 | 11.6 |
| Secondary care hospital | 196 | 18.5 |
| Tertiary hospital | 739 | 69.8 |

ICU: intensive care unit; ER: emergency room; OR: operating room; OPD: outpatient department.

The majority of HCWs (67.9%) reported managing COVID-19 patients. Additionally, 22.8% had encountered an infected family member, and 9.1% reported having been infected with laboratory-confirmed COVID-19 themselves. One-third of HCWs (33.3%) had registered for and/or received the COVID-19 vaccine. Almost all HCWs reported that they were aware of the B.1.1.7 variant (97.3%), with 36% reporting that they had sufficient knowledge of the variant. Although most participants (64.3%) were unsure whether the new mutation could cause a false negative polymerase chain reaction (PCR) result, 18.9% of them thought it would. Most of the respondents (59.8%) agreed that management of the new mutation would be similar to the current COVID-19 management guidelines. More than half of participants (53.9%) believed that the new SARS-CoV-2 mutation outbreak in foreign countries would result in subsequent lockdowns if it reached the KSA. When HCWs were asked to indicate their level of agreement on the need to impose tighter infection control measures due to the new mutation variant, a high level of agreement (M = 4.08 on a scale of 5, SD = 1.05) was found. Regarding worry about international travel within the next month, high levels of worry were found among participants, with the highest levels of worry associated with travel to the UK (M = 3.25, SD = 1.35), which was slightly higher than their worry levels about travelling abroad generally (M = 3.22, SD = 1.07) (Table 2).

**Table 2:** HCWs’ perceptions and experiences of the COVID-19 pandemic and its mutations

| Question | Answer | n | % |
| --- | --- | --- | --- |
| Have you managed COVID-19 patients? | No | 340 | 32.1 |
|  | Yes | 718 | 67.9 |
| Have any of your family members been infected with COVID-19? | No | 817 | 77.2 |
|  | Yes | 241 | 22.8 |
| Have you been previously diagnosed with PCR-confirmed COVID-19? | No | 962 | 90.9 |
|  | Yes | 96 | 9.1 |
| Have you traveled abroad in the last 3 months? | No | 998 | 94.3 |
|  | Yes | 60 | 5.7 |
| Have you received or registered to receive the COVID-19 vaccine? | Yes: I received the first dose of the COVID vaccine | 126 | 11.9 |
|  | Yes: I have registered but have not received it yet | 226 | 21.4 |
|  | No | 706 | 66.7 |
| Do you know about the new SARS-CoV-2 B.1.1.7 variant that was reported internationally this month? | Yes: I am knowledgeable about this topic | 381 | 36 |
|  | Yes: but I have limited knowledge | 649 | 61.3 |
|  | No | 28 | 2.6 |

| The COVID-19 mutation may/will result in another local lockdown in Saudi Arabia if it causes local outbreaks. | Agree | 570 | 53.9 |
| --- | --- | --- | --- |
|  | Neither agree nor disagree | 380 | 35.9 |
|  | Disagree | 108 | 10.2 |
| The mutation MAY cause a false NEGATIVE PCR result. | Agree | 200 | 18.9 |
|  | Neither agree nor disagree | 680 | 64.3 |
|  | Disagree | 178 | 16.8 |
| Question | Answer | Mean (SD) |  |
| With respect to the mutation, how much do you agree that healthcare workers should apply tighter infection control measures? | 1: Strongly disagree<br>2: Disagree<br>3: Neutral<br>4: Agree<br>5: Strongly agree | 4.08 (1.05) |  |
| In relation to the newly reported COVID-19 mutation, how worried are you about international travel? | 1: Not at all worried<br>2: Slightly worried<br>3: Moderately worried<br>4: Very worried<br>5: Extremely worried | 3.22 (1.17) |  |
| How worried would you be to travel to the following countries in the next month?* | The UK | 3.25 (1.35) |  |
|  | Other European nations | 3.10 (1.32) |  |
|  | Other world destinations outside of Europe and the UK | 2.74 (1.23) |  |

\*This question uses the same scale as the previous question.

### 3.1 HCWs’ awareness of and sources of information about the new COVID-19 mutation

HCWs’ sources of information about the mutant variant of COVID-19 are shown in Figure 1. The most common source of information was social media networks (67%), followed by the World Health Organization (WHO) website, Saudi Ministry of Health (MOH) website, and hospital or official announcements, while the U.S. Centers for Disease Control and Prevention (CDC) website was only used by 20% of HCWs. HCWs’ awareness of and knowledge about the new COVID-19 mutation was measured using eight questions (Table 3). The overall mean awareness score was 3.76 (SD: 1.23) out of 8.00.

**Table 3:** Indicators for HCWs’ awareness of/knowledge about the COVID-19 pandemic and its mutations

| Statements about Covid-19 mutation (correct answer) | Incorrectly answered n (%) | Correctly answered n (%) |
| --- | --- | --- |
| This B.1.1.7 variant was first described in the UK. (True) | 144 (13.6) | 914 (86.4) |
| The COVID virus mutation is expected. (True) | 277 (26.2) | 781 (73.8) |
| This new COVID mutation is more contagious. (True) | 249 (23.5) | 809 (76.5) |
| This new COVID mutation is causing a more severe disease. (False) | 734 (78.8) | 224 (21.2) |
| The available COVID vaccines will be effective for new virus mutations. (True) | 510 (48.2) | 548 (51.8) |
| The mutagenic virus's impact on the respiratory system is worse than the original COVID-19. (False) | 870 (82.2) | 188 (17.8) |
| To the best of your knowledge, the mutation may/will result in another wave of the pandemic. (True; uncertain considered true) | 673 (63.6) | 385 (36.4) |
| The appearance of mutagenic viruses is a sign that herd immunity is occurring. (False) | 932 (88.1) | 126 (11.9) |

**Figure 1:**
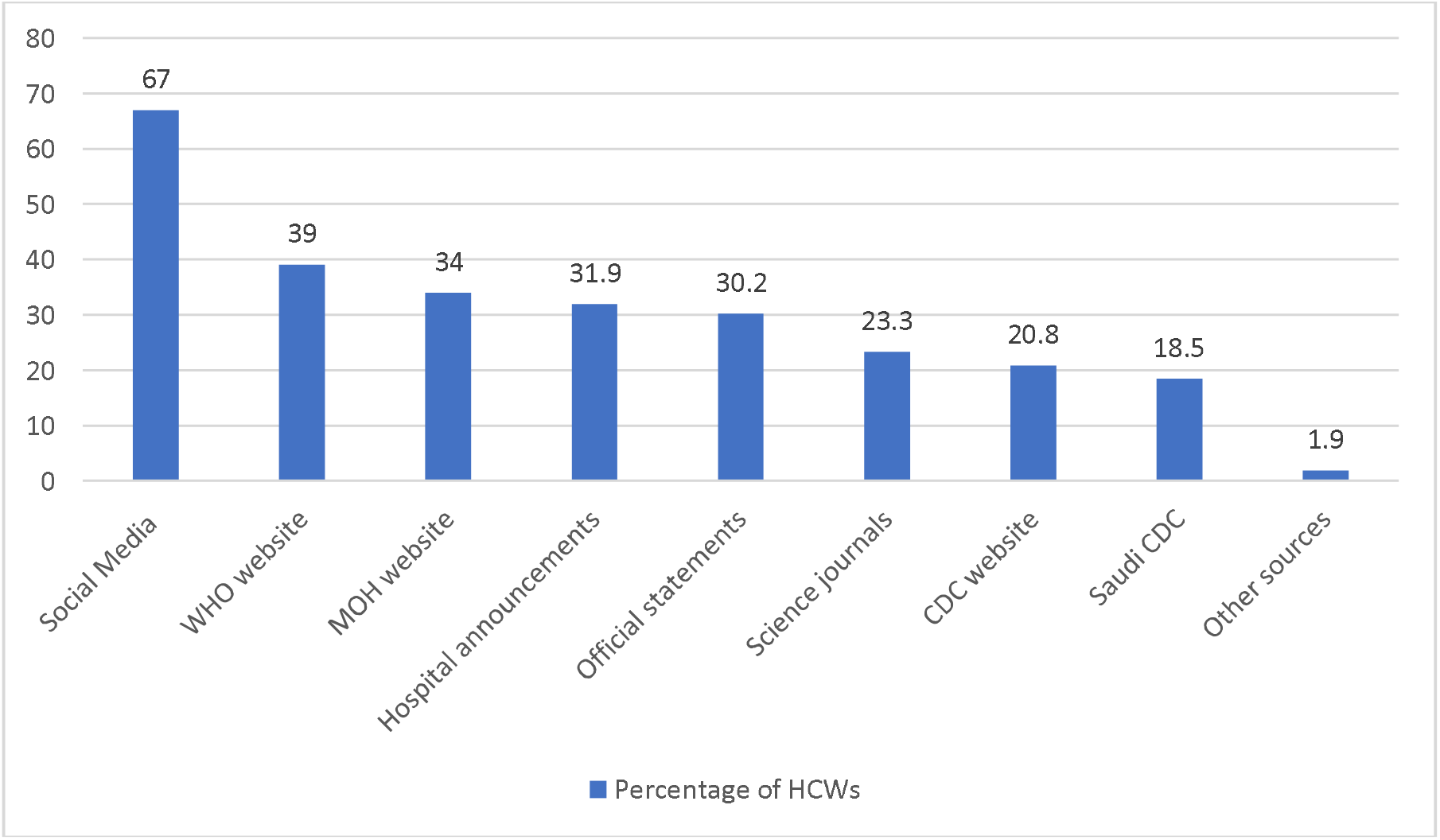
HCWs’ sources of information about the B.1.1.7 SARS-CoV-2 mutant variant.

Most HCWs (86%) were aware that the new variant was first described in the U.K., 73.8% reported that the mutation is an expected evolutionary phenomenon, and 76.5% were aware that it is more contagious. However, the majority of participants believed that the new mutation would cause a more severe disease and likely have a greater negative impact on the respiratory system. In addition, most HCWs thought that the emergence of this mutation was a sign that herd immunity is occurring.

### 3.2 International travel worries among HCWs due to the emergence of mutant COVID-19 strains

The majority of participants were expatriates living and working in Saudi Arabia. We measured HCWs’ worry levels regarding international travel. In addition, we evaluated the degree of HCW anxiety using the General Anxiety Disorder (GAD-7) scale which showed a mean score (SD)of 5(±5) for the whole group. Figure 2 showed the distribution of the degree of anxiety among HCWs.

**Figure 2:**
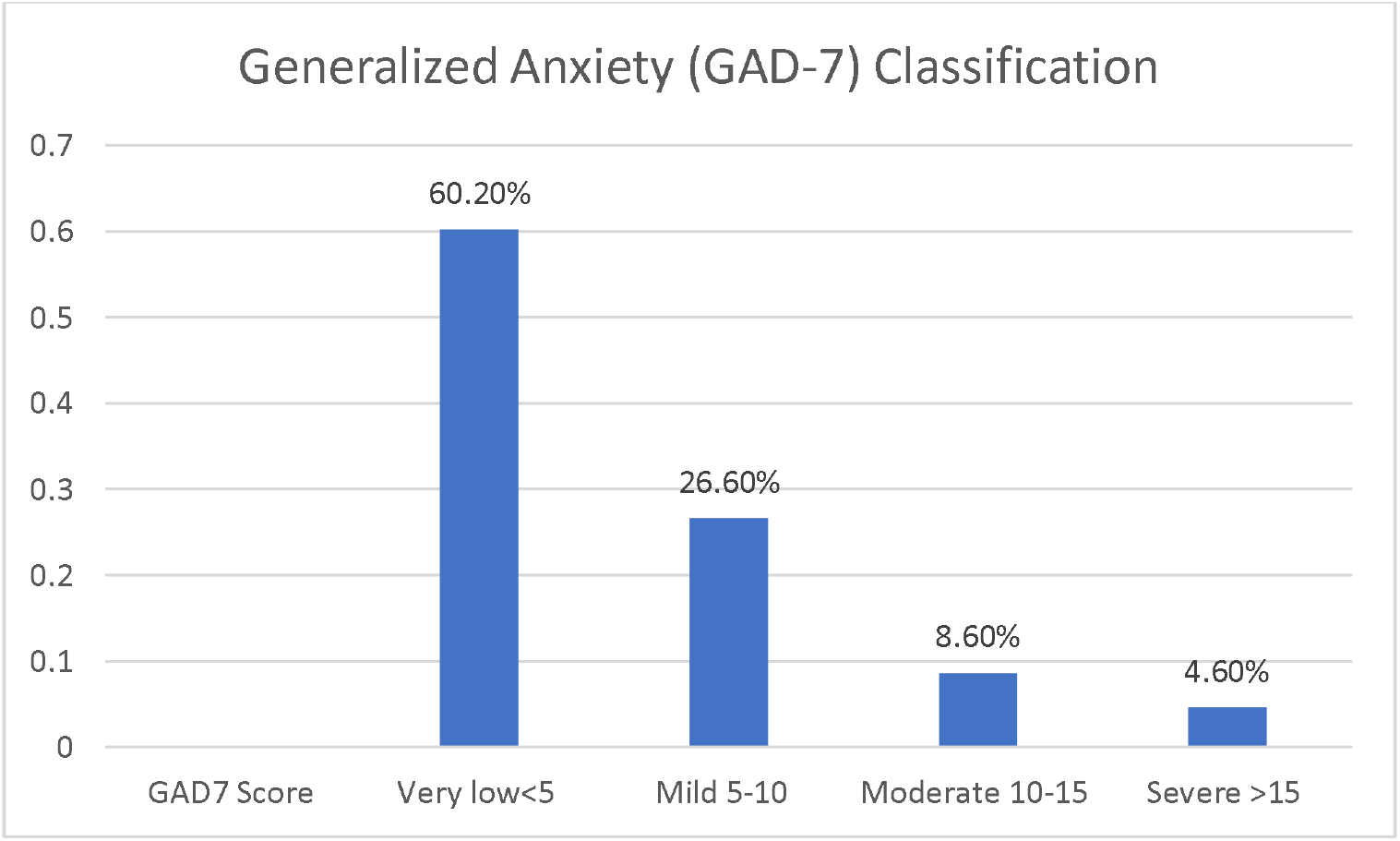
HCWs’ Generalized Anxiety Disorder Assessment (GAD-7) Score Classification

A bivariate analysis was conducted to illustrate the association between HCWs’ characteristics and their perceived levels of worry about travelling abroad due to the emergence of the new mutation (Table 4). Female HCWs (mean = 3.36, P < 0.001) and those over 50 years of age (mean = 3.55, P = 0.024) were significantly more worried about international travel than males and HCWs under 50, respectively. In addition, expatriate HCWs had a higher level of worry regarding travel compared to native Saudi HCWs (P < 0.001).

**Table 4:**
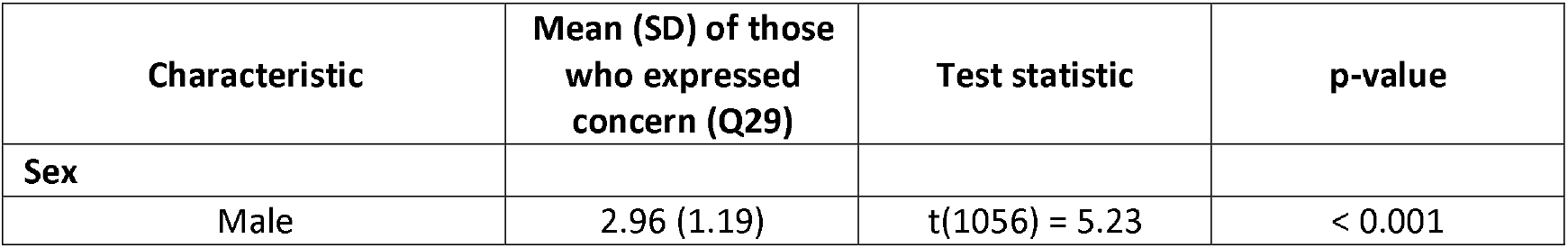
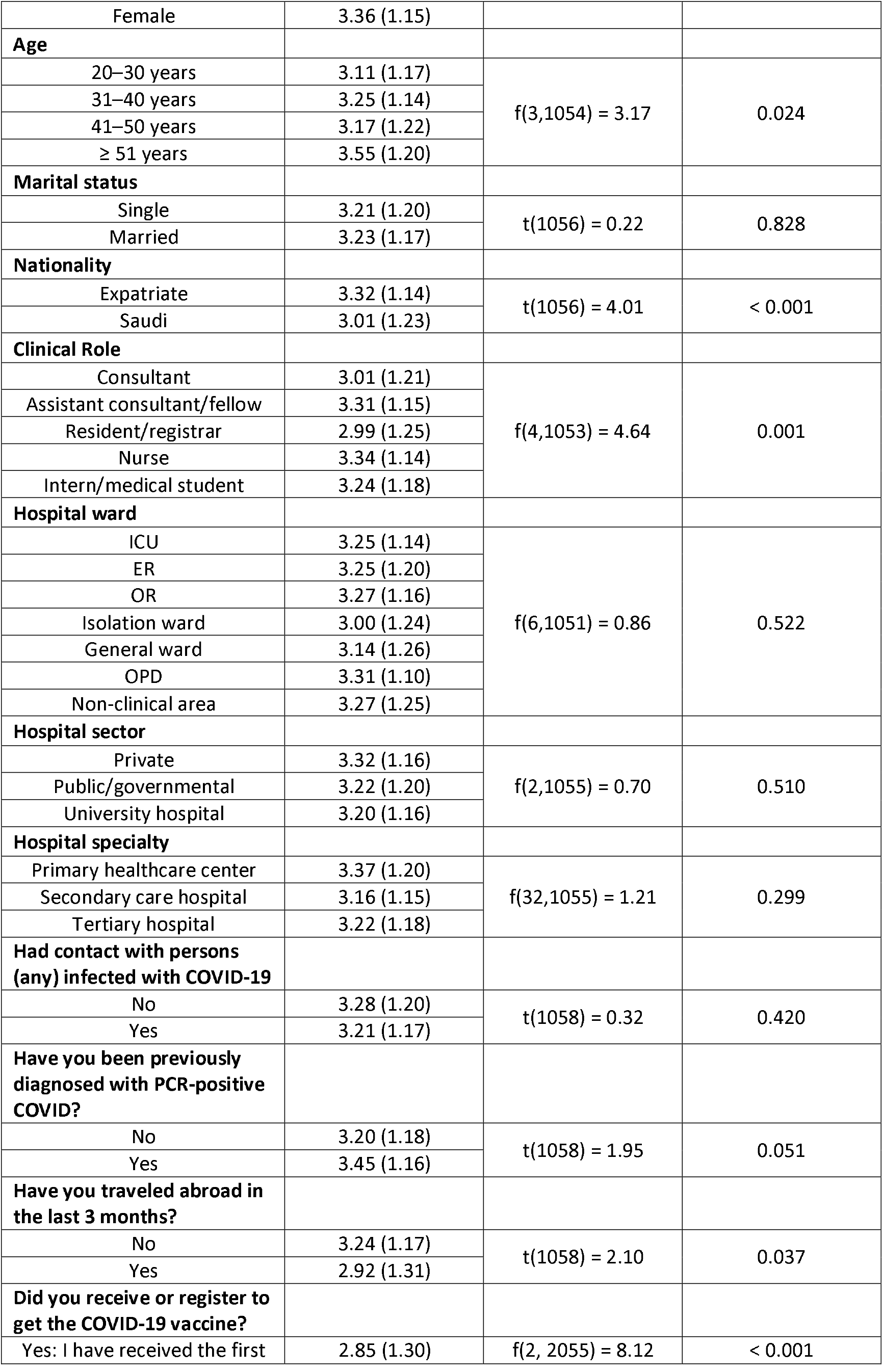
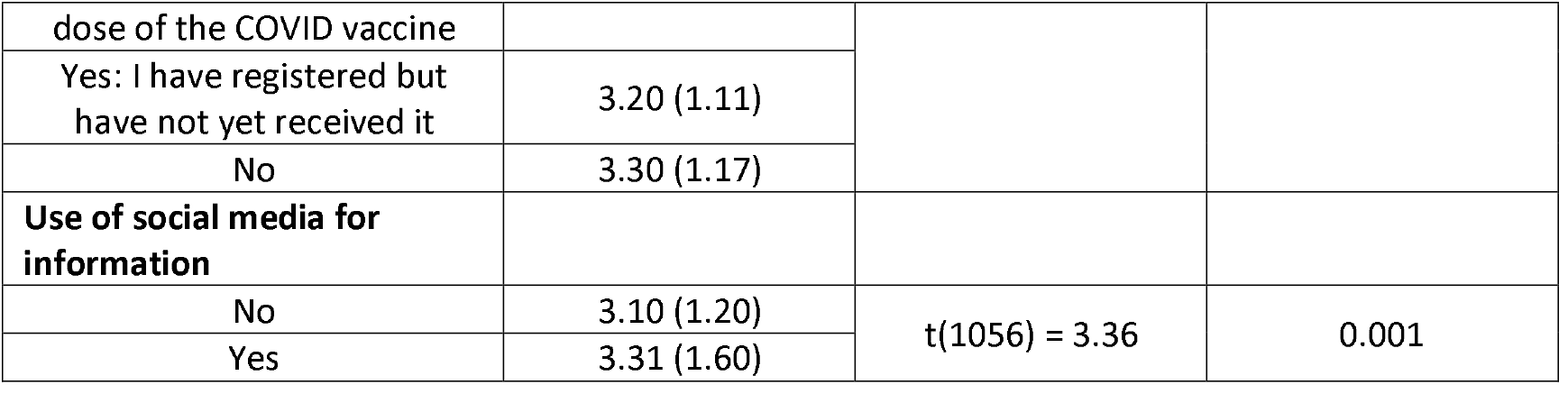
Bivariate analysis of HCWs’ characteristics and perceived worry regarding travelling abroad due to the emergence of the mutant COVID-19 strain (N = 1058)

Nurses and interns/medical students were more worried than those assigned to other clinical roles (P = 0.001). Additionally, those who reported they had not traveled in the previous 3 months and those who had not received or registered for the COVID-19 vaccine were also significantly more worried than those who had traveled or received the vaccine (P = 0.037, P < 0.001, respectively). Interestingly, those HCWs who used social media as a source of information were significantly more worried about travelling abroad due to the emergence of the new mutant strain.

Further, multivariate linear regression analyses (table 5) were used to predict respondents’ characteristics based on their level of worry associated with travelling abroad. This revealed a significant correlation for HCWs’ level of worry with older age, female sex, higher GAD-7 scores, abstinence from travelling abroad in the previous 3 months, and the belief that HCWs should apply tighter infection control measures due to the emergence of mutant strains. However, no significant correlation was found between HCWs’ level of worry and their status as having received or registered to receive the vaccine. In addition, the analysis model indicated that people who had travelled abroad recently had a significantly lower mean level of worry regarding international travel outside the UK (β = −0.437, p < 0.001).

**Table 5:** Multivariate linear regression analysis of HCWs’ level of worry regarding travelling abroad (N = 1058)

| Characteristic | Unstandardized $\beta$ Coefficient | 95% CI for $\beta$ coefficient | | p-value |
| --- | --- | --- | --- | --- |
|  |  | Lower Bound | Upper Bound |  |
| Constant | 1.483 | 0.922 | 2.043 | < 0.001 |
| Sex = Female | .208 | .037 | .379 | .017 |
| Age (years) | .018 | .010 | .026 | < 0.001 |
| Nationality = Saudi | .049 | -.121 | .219 | .575 |
| Hospital setting | -.070 | -.165 | .024 | .144 |
| Confirmed as previously having COVID-19 | .143 | -.086 | .372 | .220 |
| GAD-7 score | .065 | .052 | .078 | < 0.001 |
| Mean score for awareness of mutant virus (min = 0, max = 8 points) | -.035 | -.092 | .022 | .227 |
| Had previous contact with COVID-19 infected person(s) | -.142 | -.301 | .017 | .079 |
| Had travelled abroad in the last 3 months | -.437 | -.723 | -.152 | < 0.001 |
| Had registered/received the COVID-19 vaccine | -.058 | -.208 | .092 | .448 |
| Believes HCWs should apply tighter infection control measures | .230 | .165 | .296 | < 0.001 |
| Used social media for information on COVID-19/mutated viruses | .129 | -.015 | .272 | .080 |

Furthermore, a bivariate Pearson’s correlation test was conducted and showed that HCWs’ worry about international travel correlated significantly and positively with their GAD-7 scores (r = 0.310, p < 0.010) and perceived importance of applying stricter infection control measures (r = 0.256, p < 0.010).

## 4 Discussion

In this first reported survey on SARS-CoV-2 B.1.1.7 lineage-related perceptions and travel worry among HCWs, most respondents were aware of the emergence of the variant and had significant anxiety regarding traveling to affected countries, especially to the UK. Females and nurses expressed the highest degree of worry, and most of our study participants were nurses (59%), with two-thirds being expatriates. Saudi Arabia’s workforce relies heavily on expatriate female nurses, as only one-third of the nursing force is comprised of Saudi citizens ^12^. Due to this dependence, travel restrictions caused by the COVID-19 pandemic, and further tightening of traveling conditions with the emergence of the new variant, many of these HCWs have been stranded abroad. This may have had a negative impact on the health care system. These expatriate nurses usually reside in housing compounds with shared rooms and facilities ^13^, but during an epidemic, such arrangements can be hazardous to the health care system. A previous MERS-CoV hospital outbreak in Saudi Arabia was in part related to such accommodations ^14^. Recently, HCWs in Saudi Arabia were exempted from the travel bans. In our survey, a few HCWs indicated that they had travelled to countries with potential cases of the new variant in recent months, despite quarantine measures on returning travelers. HCWs who return from international travel could pose a potential risk of introducing emerging variants within the hospitals they serve ^15^. Previous experiences with MERS-CoV and our findings should alert public health officials to reevaluate their long-term strategy for training and empowering local frontline HCWs.

Almost two-thirds of participants had dealt with COVID-19 infected patients, 23% had a family member who had been diagnosed with the disease, and about 10% had been infected themselves. These results are similar to a previous cross-sectional survey of nearly 1,500 HCWs in Saudi Arabia, which found that 12.8% of HCWs had been infected ^10^. In that same report, two-thirds of participants were willing to receive an authorized COVID-19 vaccine when available. However, our findings of the actual implementation of the vaccine contradict this. Despite the vaccine being authorized and available free of charge in Saudi Arabia, almost two-thirds of our participants had not registered to receive the vaccine. These findings should also alert public health officials to introduce targeted vaccine campaigns among HCWs.

Most participants were aware of the new variant reported, with more than one-third indicating confidence in their knowledge. However, only one-fifth were aware that it may cause a false negative PCR result. Recently, the U.S. Food and Drug Administration ^16^ identified three different molecular techniques that may be affected by the variant and recommended that HCWs be aware of the different genetic variants of SARS-CoV-2 that may affect test results depending on the type of molecular test used to diagnose a patient. Negative results should be considered in combination with a clinical evaluation ^16^.

Our study showed that 86% of HCWs were aware that the new variant was first described in the U.K. and that many countries have subsequently reported cases. This highlights the importance of HCWs and infection control authorities keeping up to date about those countries when assessing returning travelers.

Regarding the behavior of the mutant strain, most of our HCWs knew that it is more contagious, and they incorrectly expected it to cause a more severe disease than the original strain. The new variant has been reported to be 56% more infectious ^17^. When asked whether the current COVID-19 vaccine would prevent infection, this was met with uncertainty. It is not yet scientifically proven whether any type of vaccination will be effective in preventing infection from emerging variants. In-vitro studies have shown that the BNT162b2 vaccine neutralizes SARS-CoV-2 viruses carrying N501Y mutations, but more clinical data are still needed ^18^.

In the current study, 67% of the respondents acquired their information from social media. Social media has played an important role in information accuracy as well as spreading misleading information during the COVID-19 pandemic ^19^. In a previous study from the KSA, 44.1% of participants used a social media platform such as Twitter ^19^. In our study, 73% of the respondents used either the WHO or MOH websites for information. This is an important finding that may direct plans for the dissemination of information about the pandemic.

It is interesting to note that in the bivariate analysis, HCWs’ perceived level of worry about travel was significantly associated with their use of social media as a source of information. This association may be related to the high degree of fear that tends to be reflected in the media. However, a study from the U.S. found a decreased level of concern about COVID-19 among individuals who used news outlets such as CNN or Fox News as their primary source of information ^20^.

Other factors that were significantly associated with HCWs’ decreased level of worry were being male, older age, and being a Saudi national. Several factors have been shown to contribute to people’s general levels of anxiety about COVID-19. A review study from China and India that included 18 studies with a total of 79,664 participants showed that the level of stress among the general population regarding COVID-19 was associated with having patient contact and being of the female gender ^21^. In addition, our study showed increased levels of worry among nurses and assistant consultants/fellows; however, it is important to keep in mind that the level of worry about COVID-19 is also a factor of time, as one study showed variable anxiety levels as the pandemic has evolved ^22^. It is expected that individuals who worry about additional lockdowns would also be concerned about travel, as shown by Lee et al. ^23^. It is interesting to note that in our study, participants who answered that the mutation may cause a false negative PCR result were more worried about travel. The SARS-CoV-2 mutation may or may not result in a false negative PCR result based on the location of the mutation and the molecular test being used ^24^.

COVID-19 has been associated with significant anxiety among HCWs in Saudi Arabia ^9,25^. Our descriptive Pearson’s bivariate correlations analysis showed that there was a significant correlation between worry regarding travel, the perceived importance of infection control, and generalized anxiety. This is an important finding, as a previous study found that increased resilience scores is a factor that reduces the rate of anxiety related to COVID-19 ^26^.

The link between advanced age and worry regarding travel seems expected as older people have more health concerns and are more susceptible to severe forms of infection and a higher fatality rate ^27^. To add to this, HCWs have a higher risk of catching COVID-19, with an estimated risk of three to five-fold greater than that of the general population ^28^. The literature has also shown that older HCWs are more affected by the pandemic. For example, the median age of HCWs who have died due to COVID-19 in China is 55 years ^29^. As a result, the U.S. Department of Labor recommends recognizing older age as an individual risk factor and addressing it when planning for pandemics ^30^. This background could explain why HCWs in this study recommended applying tighter infection control measures than those currently in place. This may speak to their recent experiences as frontline workers during the pandemic, as most of our respondents were registered nurses.

HCWs who had abstained from travel recently were more worried about travelling abroad. Other studies have observed a correlation between threat severity and susceptibility to the virus, which has caused travel fear and resulted in travel restrictions due to the COVID-19 outbreak ^31^. Studies have reported that similar post-disaster travel behaviors were influenced by risk perceptions and motivations ^32,33^.

We used a self-reported anxiety questionnaire (GAD-7) that is designed to assess the participants’ mental health status during the previous 2 weeks, which is well-suited to the situation of emerging news about this mutation. The GAD-7 questionnaire inquires about the overall degree to which the respondent has felt nervous, with free-floating anxiety themes ^34^. A previous study showed that HCWs had a higher prevalence of anxiety at baseline prior to the pandemic ^35^. Similarly, numerous national studies have reported an increased level of anxiety symptoms in HCWs during the COVID-19 pandemic ^36-38^. To understand this finding, we hypothesize our study likely included HCWs with high baseline anxiety levels, whose anxiety levels are now increased even more due to the SARS-CoV-2 new mutation. HCWs’ travel worries due to the new SARS-CoV-2 genetic variants should be addressed, especially because HCWs are more prone to developing mental health problems such as anxiety, depression, and substance abuse during pandemics ^39,40^. Therefore, screening and supporting expatriate or travelling HCWs could improve their mental well-being, as well as advising against HCWs’ travel except for “really needed” until the pandemic crisis is over.

### 4.1 Study limitations and strengths

This study is subject to the limitations of all cross-sectional surveys, including sampling, response, and recall biases. While this work is among the first research projects to explore travel worries among HCWs in light of the travel restrictions caused by new genetic variants in the SARS-CoV-2 virus, their experiences and perceptions are likely to change as the circumstances evolve. Furthermore, HCWs’ perceptions may differ from one country to another. Whether the HCWs receive COVID-19 vaccines themselves, and depending on which vaccine type, may further change their travel worries. Additionally, this study was conducted prior to the Saudi MOH reporting on the emergence of the B.1.1.7 variant in Saudi Arabia on January 7, 2021, which might affect HCWs’ perceptions and worry levels.

### 4.2 Conclusion

Most HCWs were aware of the emergence of the SARS-CoV-2 B.1.1.7 variant and expressed substantial travel worries in relation to it. The level of travel worries was greater among HCWs who used social media as their main source of their information, who had not registered for the COVID-19 vaccine, and who had higher GAD-7 scores. In addition, public health authorities’ utilization of official social media platforms could improve the dissemination of accurate information among HCWs regarding the virus’s evolving mutations. Targeted vaccine campaigns are warranted to ensure that HCWs are aware of the efficacy of COVID-19 vaccines towards the genetic variants of SARS-CoV-2 and to increase their vaccine uptake.

## Data Availability

All the data for this study will be made available upon reasonable request.

## Abbreviations

CDC: Centers for Disease Control and Prevention
COVID-19: Coronavirus disease 2019
GAD-7: Generalized Anxiety Disorder Assessment
SARS-CoV-2: Severe acute respiratory syndrome coronavirus 2
WHO: World Health Organization

## Ethics approval and consent to participate

The study was approved by the institutional review board of King Saud University (approval #20/0065/IRB).

